# A Case Report of a COVID-19 Infection with Positive Sputum and Negative Nasopharyngeal Rapid Antigen-Based Testing: A Practical Method to Explore Sputome

**DOI:** 10.1101/2023.01.26.23285052

**Authors:** Norberto A. Guzman, Daniel E. Guzman

**Affiliations:** Princeton Biochemicals, Inc., Princeton, New Jersey 08543, USA; Columbia University Irving Medical Center, New York, New York 10032, USA

**Keywords:** sputomics, sputome, sputum, mucus, proteoforms, lateral flow immunoassays, home-based rapid testing, COVID-19, SARS-CoV-2 virus, infectious diseases, mesophilic and thermophilic enzymes, respiratory disease biomarkers, mucolytic agents, point-of-care testing, Alcalase^®^, Triton X-100, long haulers, immobilized digestive enzymes, collection-reaction tubes

## Abstract

Current COVID-19 antigen testing is primarily carried out by obtaining a specimen via nasopharyngeal swab and performing a rapid lateral flow immunoassay (LFIA) or related immunoassays. On average, a nasopharyngeal antigen-based LFIA for COVID-19 remains positive for approximately one week from symptom onset, and levels of infectivity and duration of the symptoms may depend primarily on carrying a high viral load enough to infect others. It has been proposed that patients with long-COVID, a syndrome in which patients continue to have complications of COVID with ongoing symptoms, may have occurring viral replication, despite testing negative via rapid COVID antigen testing.

We therefore propose a modified antigen-based method that exposes hidden or masked antigenic sites of viral specimens, or lingering fragments of viral proteins, present in sputum using a home-based rapid immunoassay for COVID-19. Almost all protocols for testing were performed according to LFIA kit manufacturer’s instructions for the detection of SARS-CoV-2 nucleocapsid protein antigen, one of the most predominant proteins encoded by the SARS-CoV-2 virus. However, in challenging the manufacturer instructions, one or more digestive enzymes and a detergent were added to the collected biosamples (nasopharyngeal, oropharyngeal, saliva, buccal, gargle, and sputum); this modified procedure expose hidden or masked antigenic sites of the coronavirus or cross-reactant antigenic sites of related or non-related viruses, or some of the plethora of epitopes generated by the sample corresponding microbiota to accomplish an optimal binding to the commercial antibody used in the diagnostic test. The modified protocol can enhance detection sensitivity by making the resultant test band in sputum samples visible, that would otherwise not be seen, and consequently may generate a false negative result, in a nasopharyngeal sample from a patient with mild symptoms of COVID-19 and/or low viral load. Therefore, a need exists for an improved sample pre-treatment extraction procedure that allows optimization of sample preparation to attain a more accurate test result. Although the experiments described here were performed using commercial platforms, with antibodies directed to SARS-CoV-2 nucleocapsid antigen, this method may also be viable for the detection of any other pathogen in sputum by using antibodies directed to the key antigens present in the pathogen of interest. Furthermore, this modified method to expose the content of sputum can be used as a simple protocol to study the sputome, the proteome of sputum, and other omics (sputomics). In summary, this simple method is non-invasive, rapid, inexpensive, accurate, and may provide increased sensitivity and specificity in the detection of COVID-19 antigens for several weeks or even months.

## Introduction

Recent decades have seen repeated pathogen emergence from wild or domestic animal reservoirs to human populations, including HIV-1 and HIV-2, Middle East Respiratory Syndrome (MERS) coronavirus, and SARS-CoV-2 virus [1]. Since the beginning of 2020, the COVID-19 pandemic has become one of the most fatal pandemics in history and remains a large burden to healthcare systems globally [2,3]. Despite recent significant advances in infectious disease diagnostics, the COVID-19 pandemic has again emphasized the importance of developing point-of-care (POC) diagnostics for timely prevention and control of disease spread [4,5].

POC diagnostics or POC testing (POCT) refers to diagnosis done in the proximity of patient care that is real-time, no sample processing, rapid, accurate, and cost-effective. In the past years, with the high demand for on-site near-patient testing or bedside testing in clinical medicine, especially in resource-poor regions, POC diagnostics have attracted increasing attention since their emergences and there has been a huge market of POC devices and is an alternative diagnostic approach to laboratory analysis [5]. While this rapid format is less resource-intensive than laboratory testing, it is more subject to error; when incorrectly performed or inappropriately utilized, POCT devices may not be as reliable as laboratory-based biomarker testing as greater number of falser positive and some negative results can be obtained, that thereby require additional follow-up at an increased cost and risk to the patient. One popular POCT diagnostic method is a lateral flow immunoassay (LFIA), also known also as immunochromatographic testing. It has been successfully used for the last six decades in the detection of many diseases and conditions because it allows rapid detection of molecular ligands in biosubstrates [6]. Due to COVID-19, the LFIA platform, used for its rapid antigen test, has become more popular than ever before [7]. The LFIA technique for the early diagnosis of COVID-19 has demonstrated the ability to prevent the spread of and curtail existing COVID-19 infections. However, there is still a need for refining the LFIA for improved sensitivity and specificity in the post-pandemic era. For example, the LFIA antigen detection is based on the double-antibody sandwich method, which requires a capture antibody and detection antibody, for testing the presence of SARS-CoV-2 in biological fluids. It is often quite laborious to screen for the correct antibody pairs, and their binding need to be specific enough to avoid cross-reaction with other forms of coronavirus that have highly homologous functional domains [8]. It has previously been discussed [9] that immunoassays are prone to generate false positive results due to the polyreactivity of antibodies and antibody-like molecules. One of the most prominent causes of antibody cross-reactivity or multispecificity is molecular mimicry. Molecular mimicry is structural, functional, or immunological similarities shared between macromolecules found in infectious pathogens and in host tissues. Therefore, a false positive can result from binding a substance different from the one of interest due to structural similarities in its antigens [9].

Diagnostic tests for respiratory viral infections traditionally use nasopharyngeal samples. Sputum, on the other hand, is rarely used for viral testing [10] given its viscous nature becomes difficult to process, making its use in clinical microbiology laboratory with automated equipment impractical [11]. Mucus in the lungs is known as phlegm or sputum. Mucus is a selective barrier to particles and molecules, preventing penetration to the epithelial surface of mucosal tissues. In general, mucus is a complex hydrogel biopolymer barrier located in the airways, gastrointestinal tract, reproductive tract, and the eyes [12]. It is continuously produced, secreted, and finally digested, recycled, or discarded and its main function include lubrication of the epithelia, maintenance of a hydrated layer, exchange of gases and nutrients with the underlying epithelium, as well as acting as a barrier to pathogens and foreign substances [12].

A recent study reported that thick, gummy respiratory secretions are at the heart of severe COVID-19 [13], indicating that although tens of thousands of studies have analyzed diverse biological specimens; however, not too many studies have seriously investigated sputum samples. In fact, the composition and physical properties of these respiratory secretions are poorly understood [13]. Mucus is mainly composed of water (∼95% w/w), mucins (∼0.2 to 5% w/w), globular proteins (∼0.5% w/w), salts (∼0.5 to 1% w/w), lipids (1-2% w/w), DNA, cells, cellular debris, protective factors, and waste [12,14]. Secreted mucins are very high molecular weight gel-forming polymers, long fibrous peptides coated with a complex array of glycans, secreted by epithelial goblet cells and submucosal glands. Due to their dense glycosylation, mucins are arranged in a brush-like structure [15]. The selective permeability properties of mucus have important roles in health and disease, and changes in the structure or properties of mucus can result in various diseases [16,17]. For example, mucus pore size, thickness, chemical composition, and viscoelasticity may vary depending on the pathological condition, as well as intersubjects, suggesting a strong variability of mucus molecular structures to different environments [17]. Healthy mucus contains 3% solids, with the consistency of egg white. However, mucin hypersecretion or dysregulation of surface liquid volume may increase the concentration of solids up to 15%, resulting in viscous and elastic mucus that is not cleared. In addition, dehydrated mucus adheres more readily to the airway wall [18].

Nevertheless, with the advent of molecular methods, sputum processing for the detection of respiratory viral infections has improved. In fact, it has been found that amongst tests that are able to process sputum, sputum testing information adds approximately 11% to the diagnostic yield for the detection of many common respiratory viruses [11]. Respiratory viruses have been detected in sputum samples from patients with chronic obstructive pulmonary disease (COPD), asthma, and cystic fibrosis. Recent evidence suggests that sputum processing may even be necessary, given that certain viral pathogens such as H1N1 influenza, severe acute respiratory syndrome (SARS) coronavirus, and Middle East respiratory syndrome coronavirus (MERS-CoV) associated with the lower respiratory tract may be absent in upper airway secretions or nasopharyngeal samples [11]. A recent study in India is even recommending using sputum testing as the new mass screening method for individuals affected by COVID-19 [19]. It is evident that studies on sputum content have helped improve understanding of chronic airways disease as it can identify the presence and type of microorganism, which can indicate the severity of airways disease aiding treatment and management options [20,21].

Nonetheless, it is apparent that more studies are needed to better understand the importance of the sputum content, particularly the sputome or sputum proteome and the sputomic or sputum omics, and thereby improve diagnostic testing used for various respiratory infections. Since sputum is a complex, compact molecular polymeric hydrogel structure, it is desirable to develop a simple method to forge better accessibility to its internal content that may be sheltered, hidden, or masked within the mucus network barrier: antigenic viral proteins or other viral constituents, pathogenic microorganisms, or toxic materials. This polymeric structure is the key limitation of detection of viral proteins by lateral flow immunoassay test.

This paper therefore describes a sample pretreatment extraction procedure used in conjunction with a COVID-19 lateral flow immunoassay testing that allows the release of significant sputum content, and thus a more accurate test result. The simplicity of the assay is based on the use of at least one detergent and at least one digestive enzyme to disrupt/lyse the sputum and assay of the released components of interest using a lateral flow immunoassay platform. This improved procedure can also be used for other LFIAs, permitting respiratory viruses to be detected simultaneously with other pathogens (bacteria, fungus) in a single and highly multiplexed broad assay.

## Materials and Methods

### LFIA kits

Experiments were performed using commercially available COVID-19 at-home antigen-based rapid diagnostic tests specifically designed for the detection of SARS-CoV-2 nucleocapsid protein antigen, one of the most produced proteins of the SARS-CoV-2 virus. The kits were obtained from three different suppliers: ACON Laboratories, Inc., San Diego, California, U.S.A.; iHealth Labs, Inc., Sunnyvale, California, U.S.A.; and Roche Diagnostics, Indianapolis, Indiana, U.S.A.

### Protocol for sample collection and for LFIA testing

Nasopharyngeal samples were collected separately and independently using disposable sterile nasal swabs included in the kit provided by the suppliers. The swab was introduced into the nostril as instructed by the manufacturer protocol, followed by the insertion of the swab into a tube containing an extraction buffer carrying a surfactant. After the appropriate extraction procedure and mixing of the sample material absorbed and retained onto the swab, three to four drops of the solution removed from the squeezable plastic tube were gentle applied to the sample well of the platform or cassette (see Figure 1) of the manufacturer’s test kits, known as sample pad.

**Figure 1.**
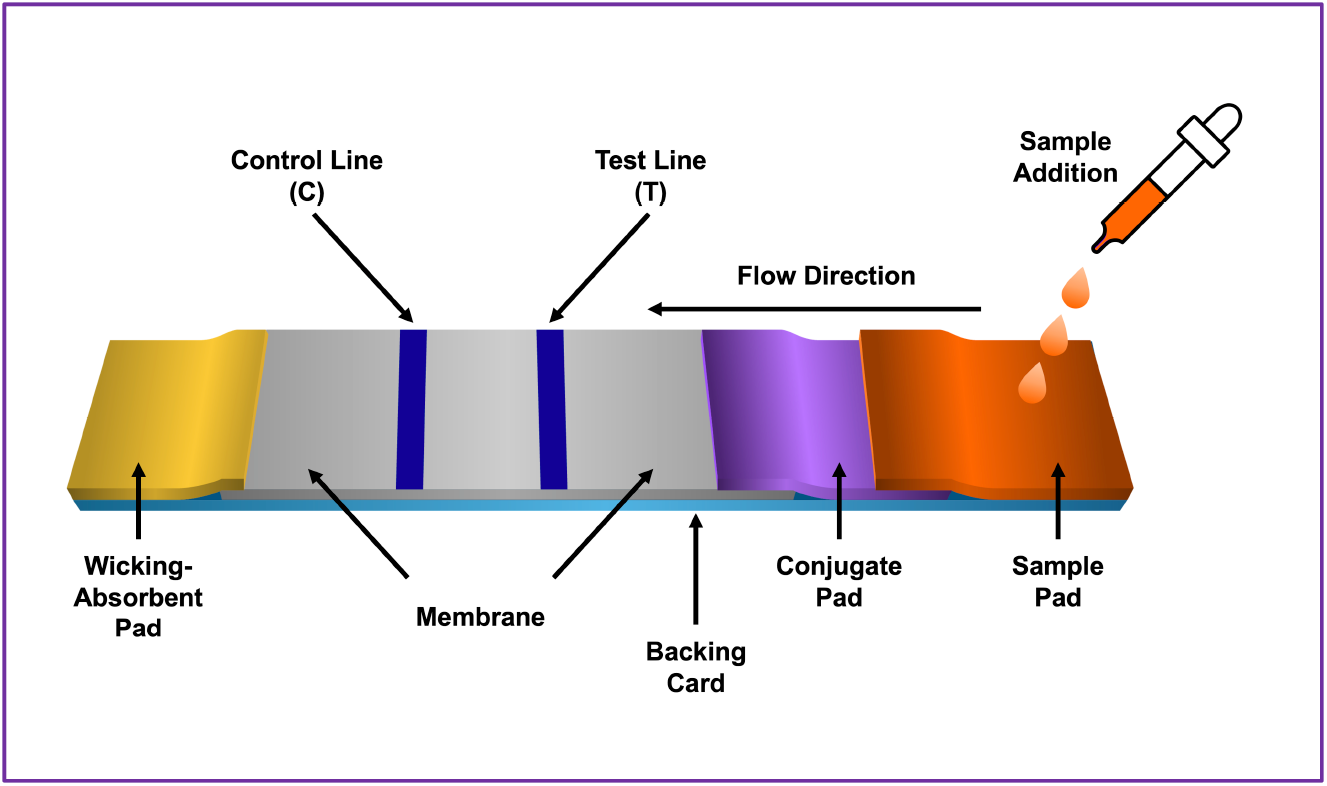
Schematic representation of a typical configuration of a lateral flow immunoassay test platform or strip. There are five key design components forming the platform: sample pad, conjugate pad, membrane with immobilized antibodies, and an absorbent pad. The components of the strip are usually fixed to an inert material backing card, housing, and protecting the test strip.

It is noteworthy to mention that the recovery efficiency and reproducibility of the sample collection depends on the material that the swab is made of and its size [22,23]. The entire procedure was carried out at a room temperature of about 25-degrees Celsius. Next, a lateral flow chromatographic migration occurs for the sample containing SARS-CoV-2 antigens to bind to the matching conjugated antibodies present in the corresponding area of the platform, known as conjugated pad. As migration occurs by capillary action, constituents of the processed-extracted sample flow progressively through the strip or platform from the sample pad to the conjugated pad, passing through the migration membrane pad and ending in the absorbent pad needed to maintain the movement. The role of the absorbent pad is to wick the excess reagents and to prevent backflow of the liquid. At the migration membrane pad or detection zone, a porous membrane (usually composed of nitrocellulose), containing immobilized antibodies in lines permits the interaction of the sample analyte (target antigen) bound to an antibody conjugated with usually micro or gold nanoparticles at the conjugated pad, resulting in the formation of a visible color. Recognition of the sample target analyte results in an appropriate response on the test line (T), carrying monoclonal antibodies, while a response on the control line (C), carrying monoclonal antibodies or polyclonal antibodies (anti-goat, anti-chicken), indicates the proper liquid flow through the strip [24]. Whether a monoclonal antibody or a polyclonal antibody is selected, it should minimally be affinity purified since any contaminating protein may compete for binding during the assay. The read-out, represented by the colored lines with different intensities, can be assessed by eye or using a dedicated reader. Depending on their size, shape, degree of aggregation, and local environment, gold nanoparticles can appear red, blue, or other colors. Gold nanoparticles (AuNP) are used as color markers with unique optical properties, extraordinary chemical stability, and binding capacity for biomolecules [25]. The control (C) line should be visible independently of the test result. When the test (T) line is visible (positive results), it indicates the presence of viral antigens in the specimen and imply that the person tested is in fact considered to be infected by the SARS-CoV-2 virus, and if no color is observed (negative results) then the person is considered not to be infected by the virus. Concerns have been raised whether rapid antigen tests for SARS-CoV-2 can result in false-positive test results [26]. Rapid antigen tests vary in sensitivity. In people with signs and symptoms of COVID-19, sensitivities are highest in the first week of illness when viral loads are higher [27]. More recently, several strategies have been implemented to enhance the signal of detection when using lateral flow immunoassays [28,29].

Other specimens, such as oropharyngeal, buccal, saline mouth rinse-gargle, and saliva were also tested, and the protocol was carried out identical to the nasopharyngeal sample using the conventional LFIA as previously described.

### Modified sample collection protocol for performing a LFIA test

Collection of expectorated sputum was carried out in collector-reactor tube and not using a swab. Collection of nasopharyngeal control samples, as well as oropharyngeal, buccal, gargle, and saliva were obtained in the same way as described above using a swab, however the processing-extraction protocol was modified before performing the LFIA test. The modified protocol consisted of adding at least one detergent followed by at least one proteolytic enzyme to the sputum sample to disperse and alter the sputum structure. The preferred detergent was Triton X-100 (Santa Cruz Biotechnology, Inc., Dallas, Texas, U.S.A.) and the preferred digestion enzyme, a protease, was Alcalase (Novozymes A/S, Bagsvaerd, Denmark). Alcalase is a versatile endoprotease providing very extensive hydrolysis. The modified extraction-lysis procedure developed for sputum was also applied to all specimens tested.

Samples of sputum were collected early in the morning, before eating or drinking, and after rinsing the mouth with clear water for about 15 seconds to eliminate any contaminant in the oral cavity as described previously [30]. After expelling saliva, the patient then breathes in deeply three times to cough at 2-minutes intervals until bringing up some spontaneous sputum. The sputum is then released in a sterile, well-closed container or vessel obtained from a local pharmacy. The sputum container can also serve as a reactor container where sputum is mixed with a detergent and a proteolytic enzyme.

About 1 mL of a Triton X-100 solution was added to approximately 2 to 3 mL of sputum. The concentration of Triton X-100 used ranged from 0.1% to 2% of total volume, of which 1% was the preferred concentration. The times of incubation at 25-degrees Celsius of the mixed sputum-detergent-enzyme solution were determined by the way the enzyme was used, either as free solution enzyme, or immobilized to a solid support, and ranged from 5 minutes to 2 hours. Regarding the quantity of Alcalase used, approximately 30-60 microliters of free-solution enzyme was added to a total volume of 3 milliliters of collected sputum sample mixed in the detergent, making a ratio of about 1-2% of enzyme-sputum solution. Other ratios were also used, depending on the viscosity of the sputum sample. After incubation, a disrupted-lysed sputum solution (see Figure 2) was decanted or centrifuged followed by the addition of 3 to 4 drops of the supernatant to the sample pad of a LFIA strip described before. The LFIA was then assayed for the presence of SARS-CoV-2 antigenic viral components. All other samples collected using a swab, the swab was immersed in a solution already containing a detergent and at least one proteolytic enzyme, following by mixing and incubation.

**Figure 2.**
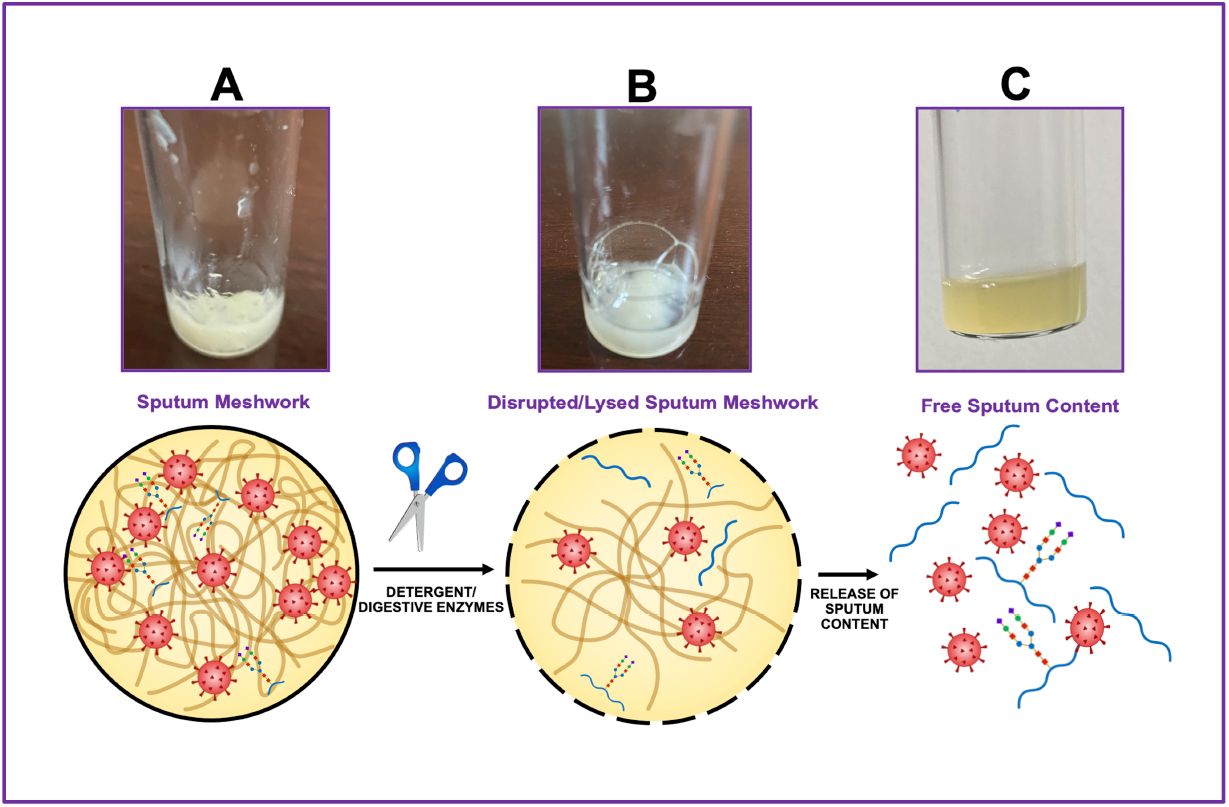
Illustrates a diagrammatic representation of the sample preparation disruption-extraction-digestion procedure applying the nonionic detergent Triton X-100 and the endoprotease Alcalase to a sputum specimen. As depicted in panel A, the sputum meshwork is a thick, rubbery, sticky, viscous, and gel-like. Sputum or mucus of the respiratory system contains numerous cells, cell debris, microorganisms, and chemical-biochemical entities. After adding the detergent and the protease some disruption occurs (panel B) influenced by time of incubation. This process resulted in a solution containing primarily soluble material and some precipitate of insoluble components (panel C). After decantation or centrifugation, the supernatant was tested for the presence of SARS-CoV-2 virus, or virus components, on a LFIA platform or strip.

### Free-solution and Immobilized Alcalase

The endoprotease Alcalase was used in the experiments reported in this paper as a free-solution enzyme and immobilized to a solid-support (Figure 3). In most experiments, the amount of expectorated sputum was abundant in the first week of performing experiments and declined significantly after several weeks. However, there was always a small amount of sputum to carried out the experiments. The proportion of sputum to detergent was maintained to an approximately ratio of 2 to 3 parts of sputum to about 1 part of detergent.

**Figure 3.**
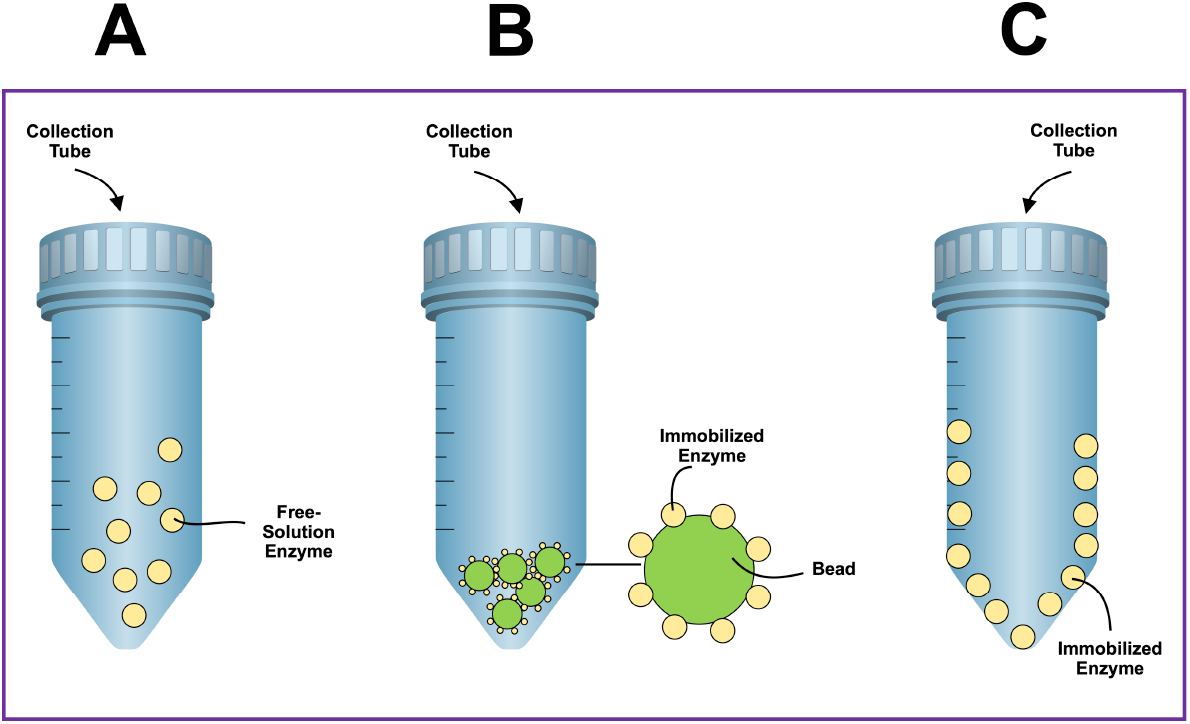
Illustrates a diagrammatic representation of collection-reaction tubes where the sputum was disrupted, extracted, and digested to release its content using the nonionic detergent Triton X-100 and the endonuclease Alcalase. The tubes feature screw caps that provide a tight secure seal. Panel A depicts the action of the protease on sputum as a free-solution enzyme in conjunction with Triton X-100. Panel B depicts the action of the protease on sputum as an immobilized enzyme to a bead used as solid support. Panel C depicts the action of the protease on sputum as an immobilized enzyme to the inner surface of a collection-reaction tube used as a solid support. The tube can be made of glass or polymeric material with a modified surface to attach one or more digestive enzymes.

The covalently immobilization of Alcalase on a solid support was carried out by previously described procedures for other enzymes or proteins [31-33]. Two methods were primarily used in our laboratory to immobilize Alcalase to beads. The first method used an epoxy activated beaded resin with a high density of epoxy-functionality (ToyoPearl AF-Epoxy-650M, Tosoh Bioscience LLC, King of Prussia, Pennsylvania, U.S.A.). The procedure for immobilization was carried out with a 50 mM sodium phosphate buffer, pH 7.0, with minor modifications of a method described elsewhere [34]. After immobilization of Alcalase to the epoxy activated resin, the remaining active groups were blocked with 3M glycine and the enzyme preparation was washed with an excess amount of phosphate-buffered saline. The second immobilization method used an amino activated beaded resin (ToyoPearl AF-Amino-650M, Tosoh Bioscience LLC, King of Prussia, Pensylvania, U.S.A.) and the linker 1,4-phenylene diisothiocyanate (PDITC, Sigma-Aldrich, St. Louis, Missouri, U.S.A.) as described elsewhere [35]. The tubes used for sample collection-reaction were made of glass. For the immobilization of Alcalase to the inner surface of the glass tube, a linking process of aminopropylate the silica-containing surface was employed using 3-aminopropyltriethoxysilane (Santa Cruz Biotechnology, Inc., Dallas, Texas, U.S.A.) prior to coupling the enzyme to the surface as described elsewhere [35].

## Results

### Modified protocol for a COVID-19 test using a conventional rapid antigen-based LFIA kit

The rationale for the research described in this paper was based on the experience of a patient looking for answers to certain symptoms. The symptoms included fatigue, minor headache, and productive cough without fever, rhinorrhea, loss of taste or smell, nausea, or vomiting. Physical examination by an emergency medical doctor at an Urgent Care Clinic and an on-site antigen based rapid LFIA test was performed, using a nasopharyngeal swab specimen; confirmatory PCR testing was also sent to a specialized clinical laboratory. The results of both, the rapid antigen test and the PCR were negative. It was concluded that the symptoms may be related to a seasonal flu rather than to COVID-19.

Given the coughing and production of sputum continued, it was of interest to test for the presence of SARS-CoV-2 virus in a sputum sample. The first step was to repeat the testing using the traditional protocol for LFIA starting the same day of the visit to the Urgent Care Clinic. As shown in Figure 4 and following the protocols of the different LFIA kits, a negative result for the presence of SARS-CoV-2 virus was obtained when testing specimens obtained with a swab from nasopharyngeal, oropharyngeal, buccal, saline mouth rinse-gargle, and saliva samples.

**Figure 4.**
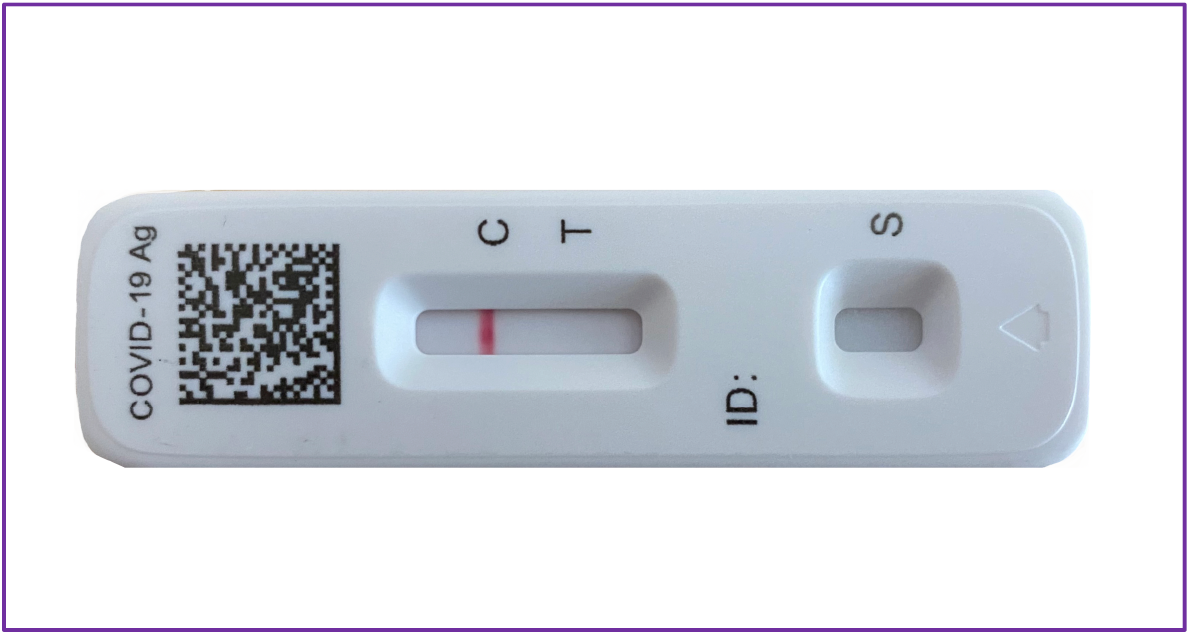
Illustrates a diagrammatic representation of a sandwich format of a lateral flow immunoassay test using the conventional protocol. When a sample containing the SARS-CoV-2 viral target is applied on a sample application pad, it flows under capillary action and color appears at test (T) and control (C) lines. However, when sample without the viral target analyte is applied on sample application pad, it flows but color appears only on control (C) line indicating a negative result. As depicted in this Figure, all tested specimens, nasopharyngeal, oropharyngeal, buccal, saline mouth rinse-gargle, and saliva samples yielded negative results. It is worth mentioning that all pictures shown in the various studies, were taken within 25 minutes from the moment the sample was applied in the sample well of the LFIA platform or strip to get the maximum color of the reaction. As times passed by the color intensity decline.

The next step was to test the same specimens but using the modified disruption-extraction detergent-protease sample preparation protocol described previously. As shown in Figure 5, a negative result for the presence of SARS-CoV-2 virus was also obtained when testing swab specimens from nasopharyngeal, oropharyngeal, buccal, saline mouth rinse-gargle, and saliva samples applied to a tube containing Triton X-100 and free-solution Alcalase. Assays for all samples were carried out using the same commercially available LFIA kits. Although the intensity of the color in each kit was slightly different when apparently having the same viral load, the results were always reproducible.

**Figure 5.**
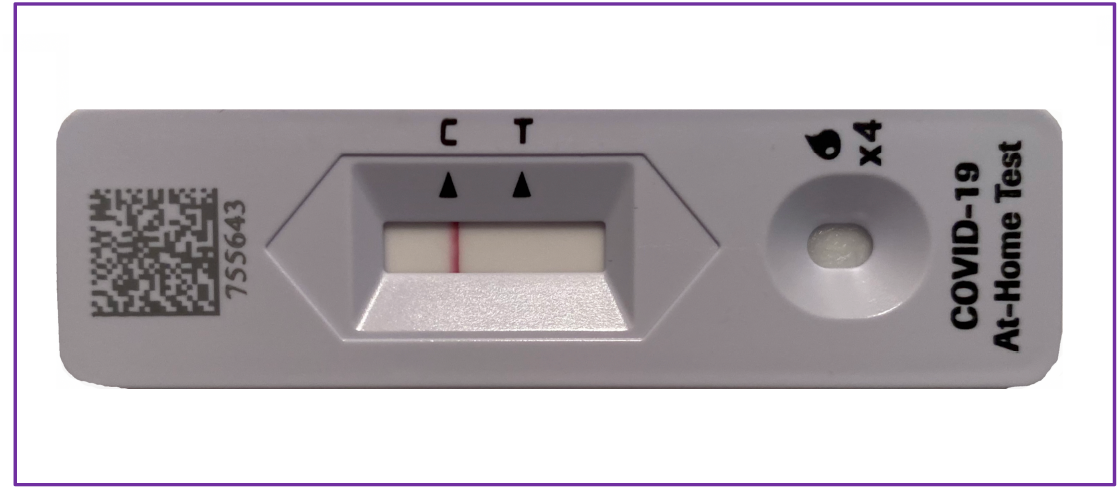
Illustrates a diagrammatic representation of a sandwich format of a lateral flow immunoassay test using the modified disruption-extraction sample preparation protocol. A distinctive color band was observed in the control (C) line, indicating a negative result for all specimens tested, nasopharyngeal, oropharyngeal, buccal, saline mouth rinse-gargle, and saliva samples when subjected to treatment with Triton X-100 and free-solution Alcalase.

Sputum was the next specimen to be tested. The modified extraction detergent-protease sample preparation protocol was mainly designed to disrupt sputum, a complex hydrogel, polymeric in composition, viscous specimen, and difficult to work with. On the first week of the manifestations of the symptoms, the production of expectorated sputum was abundant and easily to be collected but declined steadily as time passed by. Once the sputum was collected, Triton X-100 and Alcalase as free-enzyme were added to the collection-reaction tube, and the mix was incubated at 25-degrees Celsius for a period of 5 minutes to 2 hours inverting the tube a couple of times and using a constant and gentle wrist rotation motion of the tube. The tubes feature screw caps that provide a tight secure seal. After the incubation period, the disrupted-extracted-lysed sample mix was allowed to settle to remove by gravity or centrifugation some formed debris. A portion of the supernatant was placed in the sample pad of the LFIA platform or strip to let the sample migrate to the absorbent pad. As shown in Figure 6, a positive result for the presence of SARS-CoV-2 virus was obtained when sputum specimens were tested (color band in test or T line). Panel A shows the results of a sputum sample incubated with the detergent and protease for a short time, usually 5 to 10 minutes. Panel B shows the results of a sputum sample incubated with detergent and protease for a longer time, usually 1 to 2 hours. The intensity of the band seen in panel A was weak; however, the band intensity of an aliquot of the same sample increases with a longer incubation time, as shown in panel B. This indicates that time, temperature, and enzyme concentration seem to be crucial for an optimal disruption-extraction-digestion of the sputum sample to be able to release the maximum content of the constituents trapped within the complex meshwork barrier of the gel-like sputum. At present, experiments are underway to find strategies to shorten sample preparation time for LFIA testing. Similarly, several digestive enzymes and detergents are being tested for best performance within the assay. One experiment in progress is testing for the denaturation of the sputum biomolecules at higher temperatures and digestion of the complex gel-structure macromolecules with thermophilic enzymes, cleaving proteins, glycoproteins, but not nucleic acids. Our goal is to simultaneously test for antigenic viral proteins/peptides and viral RNA using the same processed sputum sample. This dual testing, using LFIA and PCR, could simplify the sample preparation procedure, avoiding the use of additional chemicals and work time.

**Figure 6.**
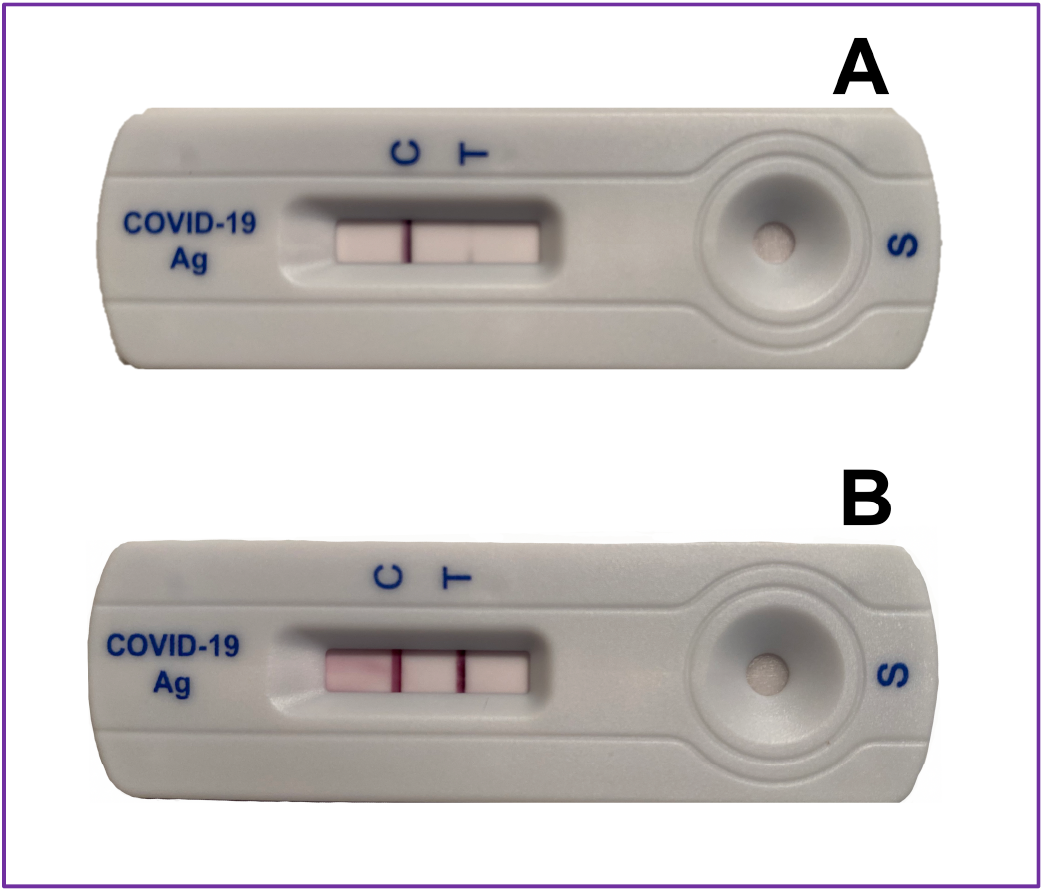
Illustrates a diagrammatic representation of a sandwich format of lateral flow immunoassay test using the modified disruption-extraction sample preparation protocol for sputum. Panel A shows an aliquot of the sputum-detergent-protease mix tested at approximately 5 minutes incubation at 25-degrees Celsius. Panel B shows an aliquot of the sample mix incubated for about 1 hour at 25-degrees Celsius.

Impressively, since the first days that some of the symptoms of COVID-19 were shown in an apparent infected person, the presence of the SARS-CoV-2 virus was reported negative using the LFIA testing for all specimens assayed, including nasopharyngeal, oropharyngeal, buccal, saline mouth rinse-gargle, and saliva, except when testing a sputum specimen in which the results were reported positive. Is the viral load in sputum detectable for 3 days, 5 days, or more, when almost all symptoms have disappeared? To answer to this question, it was necessary to employ the modified disruption-extraction sample preparation protocol and search for the presence of the SARS-CoV-2 virus in sputum samples several days beyond, when the symptoms no longer persisted.

As shown in Figure 7, a positive result was observed for the presence of SARS-CoV-2 virus tested in sputum specimens several weeks after obtaining expectorated sputum. As the frequency of coughing decreases with time, the amount of spontaneous sputum diminished as well. Nonetheless, the amount obtained for 15 weeks was sufficient to perform the experiments. To make sure all experimental conditions were consistent, strict protocols were maintained including the use of the same LFIA COVID-19 antigen-based test kits (Flowflex COVID-19 antigen home test, Acon Laboratories, Inc., San Diego, California, U.S.A.). Since the intensity of the colored band diminish in time, all tested samples were incubated for 2 hours to ensure the visibility of the band.

**Figure 7.**
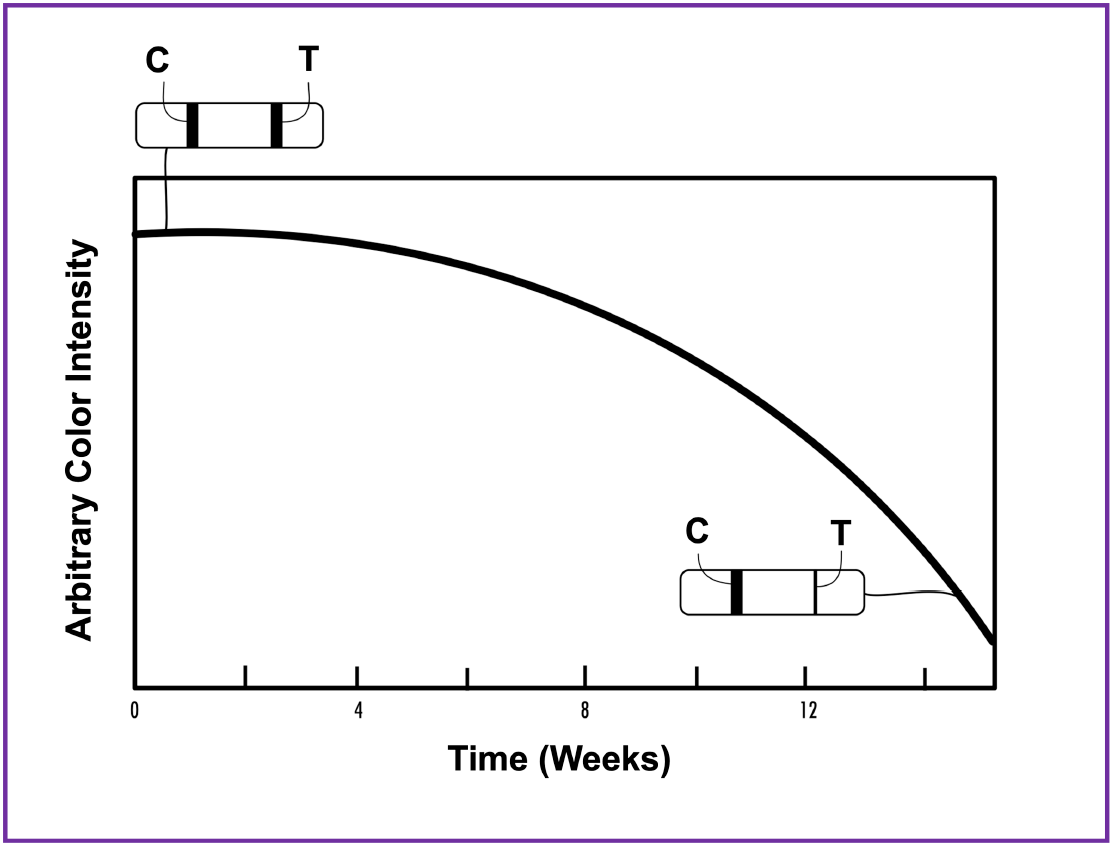
Illustrates a time experiment for the presence of SARS-CoV-2 virus in sputum samples obtained during a period of 15 weeks using the modified disruption-extraction sample preparation protocol. The color intensity of the test (T) line diminishes as the persistence of the coughing in the patient lessen, and the amount of sputum collected is reduced as well.

Preliminary experiments using the protease enzyme immobilized to the surface of a beaded resin was performed (Figure 8), and immobilization of Alcalase to the inner surface of a tube are under way. The beads used for immobilizing the protease were 65 microns, limiting the surface area for linking the proteolytic enzyme. Nonetheless, the procedure works demonstrating the presence of viral entities in the sputum. Increasing the surface area for enzyme immobilization, using smaller bead size, may enhance color band intensity.

**Figure 8.**
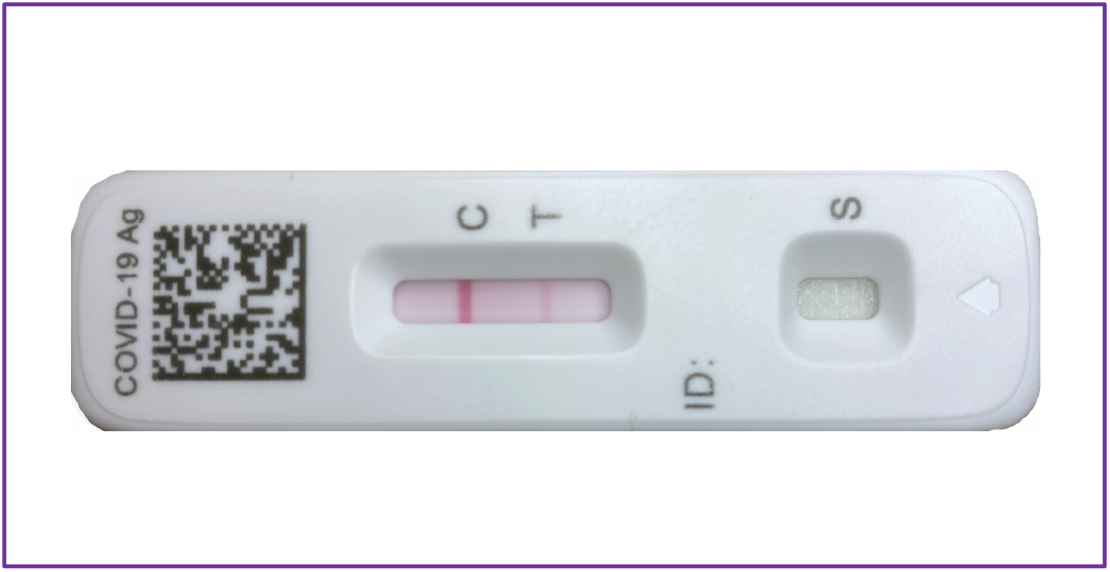
Illustrates a diagrammatic representation of a sandwich format of lateral flow immunoassay test using the modified disruption-extraction sample preparation protocol for sputum, but immobilized Alcalase to a beaded resin was used. Incubation of the sputum in Triton X-100 with the immobilized protease was 2 hours at 25-degrees Celsius.

## Discussion

Clinical diagnostic sputum tests aim to detect the causes of lower respiratory tract infections and some other diseases. The tests also provide an efficacious tool for monitoring the effectiveness of clinical treatment [30]. However, the usefulness of sputum culture in guiding microbiological diagnosis in some infectious diseases is controversial [36,37]. For example, in the case of tuberculosis, the infectious nature, prevention and control of tuberculosis has remained a topic of controversy and unresolved issues for over 1,000 years [38,39]. At the present time, tests that can replace sputum smear microscopy have been identified as a top priority diagnostic need for tuberculosis by the World Health Organization [40]. Tuberculosis is the 13^th^ leading cause of death and the second leading infectious killer after COVID-19 (above HIV/AIDS). More than a third of people infected with tuberculosis went undiagnosed in 2020, with each case infecting 15 more [41].

Similarly, with the growing crisis of the coronavirus disease (COVID-19), companies, universities, research institutions, and investigators over the world are looking for several approaches to address the challenges of the SARS-CoV-2 virus, to mitigate the spread and develop a cure for this disease. To curb the spread of the COVID-19 pandemic, the world needs diagnostic systems capable of rapid detection and quantification of the novel coronavirus SARS-CoV-2. At present, molecular techniques based on the real-time reverse transcriptase-polymerase chain reaction (RT-PCR) are considered the gold standard for COVID-19 diagnosis among other diagnostic methods. However, the RT-PCR test for SARS-CoV-2 virus does have some pitfalls that necessitate improvements in the way the method is used [42]. As with immunodiagnostic tests, the RT-PCR test can have difficulties in distinguishing between true positive and true negative COVID-19 infected individuals. It is a wise precaution not to rely on PCR test results alone, and to consider other clinical and molecular evidence [43]. Erroneous positive and negative molecular diagnostic results are indeed important and can have serious implications in people. Therefore, the use of RT-PCR combined with immunodiagnostic tests, such as LFIA or ELISA (enzyme-linked immunosorbent assay), can enhance the overall sensitivity and reduce erroneous information [44-48].

As clearly demonstrated in this paper, a rapid antigen-based lateral flow immunoassay test performed at an Urgent Care Clinic and a PCR test for the detection of SARS-CoV-2 virus in a nasopharyngeal swab specimen, resulted in a negative diagnosis for COVID-19. However, a search for SARS-CoV-2 in a sputum specimen tested positive using a rapid antigen-based LFIA platform. Should the erroneous results reported by the Clinic be considered a false diagnosis? Should the patient be considered a silent carrier of the virus without knowing? Could the carrier individual be infecting others, since the positivity of the antigen based LFIA test for sputum lasted at least 15 weeks? Particularly when symptoms disappeared after 4 days. Could this condition be transformed into what is termed post-COVID condition or long-haulers? Could findings from patients with negative pharyngeal swabs, using rapid antigens tests and PCR testing, be used as the sole basis for infection control decisions, or sampling of additional body fluids should be considered? Should a PCR test be necessary for a sputum sample, when the antigen based LFIA yields positive results for a period of 15 weeks, and the nasopharyngeal test turns negative after the first week? A fundamental aspect to be considered is that the antigen based LFIA results are either positive, weakly positive, or negative, and if repeat testing is required, the assay is inexpensive when compared to an inconclusive or invalid PCR result in need of a retake test.

It is known that in SARS-CoV-2 viral RNA is still present in feces of more than 60% of patients, after nasopharyngeal swab testing turned negative by RT-PCR assays, suggesting that fecal-oral transmission may serve as an alternative route for SARS-CoV-2 transmission [49-51]. Other studies found at day 111 from a COVID-19 patient, that in specimens from a nasopharyngeal swab and sputum inoculated into VeroE6/TMPRSS2 cells, cytopathic effects were observed, and viral RNA was detected in the culture supernatant by quantitative RT-PCR [52]. The Centers for Disease Control and Prevention recommend that patients infected within the past 90 days without new COVID-19 symptoms should not be retested [53]. Can people with no symptoms and that have not been tested for COVID-19 in sputum still be carriers of the virus and be potential infectors of others? Many questions are still unanswered, and controversies have been reported related to the COVID-19 disease, its treatments, secondary effects due to vaccines and vaccine hesitancy, and even about the accuracy and performance of the diagnostic tests currently available [54-58].

There are several advantages for treating a sputum specimen with the nonionic detergent Triton X-100 and the protease Alcalase. It can disrupt, extract, and digest at least partially the complex sputum meshwork to generate enough antigenic virus and/or viral particles to produce a positive result for COVID-19 using a rapid antigen test. Certainly, other microorganisms, chemical and biochemical entities might also be released from the sputum as well. With the use of immobilized digestive enzymes (Alcalase^®^, Flavourzyme^®^, Protamex^®^, Neutrase^®^, trypsin, pepsin, amylase, hyaluronidase, and/or other digestive enzymes) onto nanoparticles, and of detergents (Triton X-100, Nonidet P-40, and other surfactants), the release of virus and/or viral constituents can be faster due to a larger quantity of digestive enzymes present in the sample preparation. Additionally, pre-packed tubes or containers carrying detergents and immobilized enzymes, can be used as a sample collector, reactor, and as a sample transporter tube as well, since positive results can be observed in a LFIA strip after sputum specimen interaction with Alcalase and detergent for at least 4 days at 25-degrees Celsius, albeit with a slight decrease in color intensity.

Spontaneous expectoration of sputum or induced sputum can be simple to obtain, and the sample processing method using detergents and digestive enzymes to test for SARS-CoV-2 virus is practical and cost-effective, when compared to PCR tests that requires trained personnel and an equipped laboratory. Furthermore, the release of sputum constituents using this protocol can be of utility to study Sputomics (all omics in sputum) and Sputome (all proteins in sputum). The information of sputum in normal and disease state could be so vast that it may be considered the barometer of health. Studies of proteins in sputum have been carried out by several laboratories and more than 600 proteins found in sputum can be considered possible biomarkers of diseases [59-62]. Detection of sputum transferrin and urease has been found to be highly associated with pulmonary tuberculosis infection [59]. Additionally, the use of Alcalase immobilized to nanospheres may have a potential as a mucolytic agent. While mucus can be beneficial to the body, producing too much mucus very likely may cause breathing difficulties and infection [63]. Muco-obstructive respiratory diseases, including chronic obstructive pulmonary disease, asthma, and cystic fibrosis are characterized by airways obstruction due to thick, adherent mucus. Reducing the viscoelasticity of airway mucus is an area of constant research [64]. Also, nanoparticle-based delivery systems are becoming promising for treatment of respiratory treatment [65]. Advancement in nanotechnology using hybrid nanoparticles in the range of 200 nanometers have provided significant impetus to inhalation drug delivery, particularly in the management of pulmonary aspergillosis infection [66].

Over the last century, outbreaks and pandemics have occurred with disturbing regularity. The emergence and spread of pathogens will continue to be a part of our lives, as new variants of diverse pathogens will persist through time. Technological surveillance systems should be established with simple, rapid, accurate, and inexpensive assays. Advancements in finding effective therapies and vaccines should also be hand-in-hand with new and error-free diagnostic assessment. Although point-of-care tests are the most desirable alternative to ramp up large-scale population screening, analytical separation technologies coupled to selective affinity-capture methods and powerful detectors, such as laser-induced fluorescence and mass spectrometry are becoming vital tools for isolation, separation, identification, and structural characterization of molecular viral and other pathogens entities [9, 67-73]. Infectious diseases do not respect international boundaries. Consequently, all countries should join and support the efforts needed to achieve global control and prevention of large-scale outbreaks of infectious diseases [74,75].

The limitations of our study are that is based on a single patient, although serial samples were obtained for 15 weeks, showing consistency of results. Still these results warrant further studies in large scale to make this important observation clinically significant.

## Conclusion

A simple, rapid, and cost-effective method to study the constituents of sputum is described. Using commercially available lateral flow immunoassay kits, and inexpensive reagents such as detergents and proteolytic enzymes, it was possible to challenge a negative nasopharyngeal rapid antigen-based testing for COVID-19 with a positive testing. This method may simplify the current tedious protocols using strong and hazardous chemicals to extract biomolecules and to breakdown complex structures, such as dithiothreitol, guanidinium isothiocyanate, phenol-chloroform, and others, as well as the use of silica-based columns. Pre-packet collection-reaction tubes with immobilized Alcalase enzyme and detergent would be a simpler, safer, and distinctly efficient method to disrupt the complex sputum meshwork and yield a more accurate test for SARS-CoV-2 viral entities.

## Data Availability

Data Availability Statement
All data generated or analyzed during the study are included in the article

## Author contributions

N.A.G. conceived, designed, performed the experiments, and wrote the paper; D.E.G. analyzed data and critically revised the manuscript. The authors have read and agreed with the final version of the manuscript.

## Compliance with ethical standards

Biological samples were obtained from one volunteer from the start of the symptoms for 15 weeks with written and informed consent. The dated and signed consent document has been archived.

## Safety Statement

No unexpected or significant safety hazards are associated with the reported work.

## Data Availability Statement

All data generated or analyzed during the study are included in the article.

